# Rising Trends in Diabetes and Fasting Plasma Glucose among Bangladeshi Adults: Insights from National Surveys (2011–2022)

**DOI:** 10.1101/2024.10.30.24316405

**Authors:** Juwel Rana, John C. Oldroyd, Mohammad Bellal Hossain, Sadia Katha, Md. Nuruzzaman Khan, Md. Nazmul Karim, Stefano Renzetti, Rakibul M. Islam

## Abstract

**Background:** Bangladesh faces a rapidly increasing diabetes burden; however, current evidence is lacking. This study examines recent trends and identifies risk factors associated with changes in the prevalence of diabetes and fasting plasma glucose (FPG) among adults in Bangladesh between 2011 and 2022.

**Methods:** Data from three cycles of the Bangladesh Demographic and Health Surveys from 2011 to 2022 were analysed, involving individuals aged 35 years and older. Diabetes was defined according to the WHO criteria as a FPG level of 7.0 mmol/L or higher, and/or self-reported use of glucose-lowering medication. Age-standardised diabetes prevalence was calculated using the direct method, and trends were examined using a generalised estimating equation and generalized additive models.

**Results:** The overall age-standardised prevalence of diabetes in 2022 was 20.4% (95%CI: 19.2-21.7), significantly rising from 10.7% (95%CI: 9.8-11.6) in 2011 and 12.8% (95%CI: 11.7-14.0) in 2017-18. After adjusting for potential covariates, prevalence ratios (PRs) increased by 37% (PR: 1.37; 95%CI: 1.31-1.43) per BDHS cycle, with women showing the largest increase (PR: 1.43; 95%CI: 1.34-1.52). Likewise, FPG levels increased by 0.32 mmol/L overall (95%CI: 0.29-0.35), with a larger increase observed in women (0.36 mmol/L; 95%CI: 0.31-0.41) compared to men (0.27 mmol/L; 95%CI: 0.23-0.32). Body mass index and socio-economic status were the key risk factors for changes in diabetes and elevated FPG.

**Interpretation:** The prevalence of diabetes and FPG levels have significantly increased among adults in Bangladesh from 2011 to 2022, especially among women. Population-level interventions are urgently needed to control this increasing burden of diabetes in Bangladesh.

**Funding:** None.

**Research in Context:** *Evidence before this study:* Diabetes prevalence is escalating globally, with the largest increases projected for low- and middle-income countries (LMICs) like Bangladesh. However, recent data on diabetes trends and risk factors in Bangladesh remain sparse, particularly across nationally representative samples. Prior studies have shown an increase in diabetes but have not captured recent trends or provided a comprehensive analysis of risk factors across a decade.

*Added value of this study:* This study offers the most up-to-date examination of diabetes prevalence and fasting plasma glucose (FPG) levels among adults in Bangladesh from 2011 to 2022, based on three nationally representative survey cycles. It reveals significant increases in diabetes prevalence and FPG, with notable gender disparities and a sharp rise in recent years, particularly among women. By identifying body mass index and socio-economic status as key risk factors, this study highlights areas for targeted interventions.

*Implications of all the available evidence:* Our findings indicate an urgent need for population-level interventions to address the diabetes epidemic in Bangladesh, particularly as diabetes rates are projected to continue rising. This evidence can guide policymakers to prioritize resources toward diabetes prevention and control efforts, especially among women, who are experiencing a disproportionate increase in diabetes risk.

## Introduction

Diabetes mellitus contributes significantly to the global burden of disease ^1^. The International Diabetes Federation (IDF) estimated that 537 million adults had diabetes worldwide in 2021, with a projected increase to 783 million by 2045 ^2^. The largest increases in diabetes burden moving forward are expected to occur in countries undergoing economic transition from low- to middle-income status ^2,3^. Currently, ∼80% of those with diabetes live in low- and middle-income countries (LMICs) ^2^. Southeast Asian countries will be particularly affected as it is expected that diabetes cases will increase by 68% in the next two decades, from 90 million in 2021 to 151 million by 2045 ^2^.

Bangladesh is a South Asian country with increasing population density and life expectancy ^4,5^. In 2021, an estimated 13.1 million adults in Bangladesh were living with diabetes, and this number is projected to nearly double to 22.3 million by 2045 ^2^. The implications of such a large burden for a country with limited resources for diabetes management are serious. For example, the direct and indirect costs of diabetes and it’s complications alone in Bangladesh are conservatively estimated to be USD 1.5 billion annually ^6^. Health expenditure attributable to diabetes care will increase if no effective preventive measures are adopted ^7^.

Despite the evidence for a high burden of diabetes in Bangladesh, recent trends in diabetes prevalence have not been examined in detail in the country. There is evidence that the prevalence of diabetes among adults has increased from ∼5% in 2001 to ∼14% in 2017 ^8–10^. However, no study has yet investigated trends in diabetes prevalence among Bangladeshi adults using recent, large, nationally representative surveys. Utilizing three cycles of the Bangladesh Demographic and Health Surveys (BDHS) for 2011, 2017-18, and 2022, we aimed to examine trends and identify risk factors associated with changes in the prevalence of diabetes and fasting plasma glucose (FPG) among Bangladeshi adults aged 35 years and older.

## Methods

### Study design and sample

The study utilized data from three cycles of the BDHS, conducted in 2011, 2017/18, and 2022 by the National Institute of Population Research and Training (NIPORT) under the Ministry of Health and Family Welfare, Bangladesh. ICF International provided technical assistance as part of the Demographic and Health Survey Program, with financial support from the US Agency for International Development. The surveys employed two-stage stratified sampling of households, which were nationally representative and stratified by rural, urban, and city corporation areas. The sampling frame used the 293,579 enumeration areas (EAs) from the 2011 national population and housing census, prepared by the Bangladesh Bureau of Statistics. In the first stage, 600 EAs were selected for the 2011 survey, and 675 EAs were selected for 2017/18 (672 EAs were included) and 2022 (674 EAs were included) surveys, with probability proportional to EA size. Each EA contained an average of 120 households. In the second stage, systematic random sampling was used to select 30 households per EA for the 2011 and 2017/18 surveys, and 45 households per EA for the 2022 survey, to provide statistically reliable estimates of health outcomes for both urban and rural areas as well as the country as a whole. This resulted in a sample of 17,964 eligible households for the 2011 survey, 20,160 for the 2017/18 survey, and 30,330 for the 2022 survey. The household-level response rates were 97.9%, 99.4%, and 99.6% for the 2011, 2017/18, and 2022 surveys, respectively. Eligible respondents in these households were ever-married women of reproductive age who had spent the night before the survey in the selected households.

Additionally, subsampling was conducted for biomarker collection, including blood glucose and blood pressure measurements. One-third of the households in each EA were selected for biomarker data in the 2011 and 2022 surveys, and one-fourth of the households in the 2017/18 survey. Biomarker data were collected from women and men aged 35 or older in the 2011 survey and those aged 18 or older in the 2017/18 and 2022 surveys. This resulted in 8,835 eligible respondents (4,311 women and 4,524 men) for the 2011 survey, 14,704 respondents (8,013 women and 6,691 men) for the 2017/18 survey, and 15,009 respondents (8,156 women and 6,853 men) for the 2022 survey. Detailed information on survey sampling and data collection procedures has been previously published ^9,11,12^. The surveys, including biomarker measurements, were approved by institutional review boards at ICF and the Bangladesh Medical Research Council. Informed consent was obtained from all participants.

### Analytical sample

We analysed data from a total of 22,359 respondents (unweighted n=22,382), focusing on individuals aged 35 years and older, after excluding those with missing data in outcomes and covariates, as well as women with gestational diabetes and hypertension. This included 7,540 respondents (unweighted n=7,562) from the 2011 survey, 6,560 (unweighted n=6,697) from the 2017-18 survey, and 8,259 (unweighted n=8,123) from the 2022 BDHS survey. The selection criteria were applied to ensure consistency in age groups across the surveys, including only those with complete data (< 1% missing) for analysis.

### Outcome measures

The primary outcome was diabetes, assessed based on fasting blood glucose levels. Blood glucose was measured using the HemoCue Glucose 201 RT analyser in the 2022 and 2017/18 surveys, and the HemoCue 201+ analyzer in the 2011 survey. In all cases, capillary whole blood was obtained from the middle or ring finger after participants had fasted overnight. These systems automatically converted fasting whole blood glucose measurements to FPG equivalent values ^9,11,12^. Diabetes was defined according to the World Health Organization (WHO) criteria as a FPG level of 7.0 mmol/L or higher, and/or self-reported use of glucose-lowering medication ^13^.

### Risk factors

Relevant literature on diabetes in Bangladesh and other South Asian countries were reviewed ^8,10,14–16^. Participants’ age, sex, education, marital status, body mass index (BMI), hypertension, wealth quintile, place of residence (urban vs rural), and region of residence (administrative divisions) were included in the model as risk factors. The presence of hypertension was defined as a systolic blood pressure (SBP) ≥ 140 mmHg or a diastolic blood pressure (DBP) ≥ 90 mmHg or current treatment with antihypertensive medication. Socioeconomic status was derived from the household wealth index reported in the BDHS. It was constructed using principal component analysis from household’s durable and non-durable assets (e.g., televisions, bicycles, sources of drinking water, sanitation facilities and construction materials) ^17^ and expressed as wealth quintile (poorest to richest).

### Statistical analysis

Descriptive data were presented with median (first and third quartiles) for continuous variables while counts and percentages were reported for categorical variables. To compare BDHS 2011, BDHS 2017-18, and BDHS 2022 surveys, t-test or Wilcoxon rank-sum (Mann-Whitney U) test and Chi-squared test or test of proportion were used for continuous and categorical data, respectively. The prevalence of diabetes was estimated by adjusting the sampling weight and complex survey design. To account for different age distributions between groups and over time, we calculated prevalence estimates age-standardised to the 2022 Census population of Bangladesh using the direct method, where k is the number of age groups, r is the prevalence of diabetes in age group l and P is the population size in the l^th^ age group. The unadjusted trends in prevalence estimates of diabetes from 2011 to 2022 were presented as a relative change.

To examine the changes in diabetes prevalence and FPG levels from 2011 to 2022, we utilized Generalized Estimating Equations (GEE) models. For diabetes prevalence, we applied a Poisson distribution with a log link function, while for FPG levels, we used a Gaussian distribution with an identity link function. An “exchangeable” covariance matrix was specified, and cluster numbers within each survey cycle were considered as the identification variable. We used the Poisson family to avoid the problem of overestimation when odds ratio is estimated using logistic regression from a common binary outcome (>=10% prevalence) ^18^. We chose GEE models because our data are clustered by survey cycles and within clusters (sampling units), participants may share similar characteristics that introduce intra-cluster correlations^19^. GEE effectively handles these correlations without any distributional assumption, providing robust standard errors and accurate population-averaged estimates of changes in diabetes prevalence and FPG levels over time. Thus, it is well-suited to infer changes in the outcomes over time (linear trends treating the time as a continuous variable) and assess risk factors associated with changes in diabetes outcomes across BDHS cycles. In addition, non-linear trends in diabetes outcomes over time, adjusting for all covariates and accounting for clustering effects, were examined using generalized additive models (GAM) with a cubic spline (knots = 3). To understand sex-disparities in the changes in diabetes over time, we tested for heterogeneity using Cochran’s Q-test statistic^20^. Survey weights were also incorporated in all analyses to account for the complex survey design. All statistical tests were two-sided, and a p-value <0.05 was considered statistically significant. All statistical analyses were performed using R 4.4.4 (R Foundation for Statistical Computing, Vienna, Austria). The study was designed and reported in accordance with Strengthening the Reporting of Observational Studies in Epidemiology (STROBE) guidelines21.

## Results

### Demographic characteristics

The analysis included a total of 22,359 participants, 7,540 from 2011, 6,560 from 2017-18, and 8,259 from the 2022 BDHS survey, with a median (IQR) age of 49.0 years (40.0, 60.0) (p=0.2). Participants from 2022 had significantly higher rates of overweight/obesity (p < 0.001), education (p < 0.001), and hypertension (p < 0.001) compared to the 2011 and 2017/18 surveys. However, there were no significant differences in sex distribution and wealth quintiles across the survey cycles (Table 1).

**Table 1:**
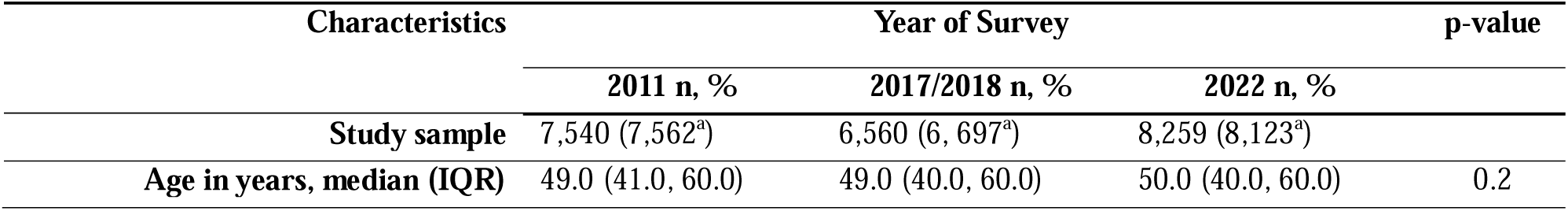
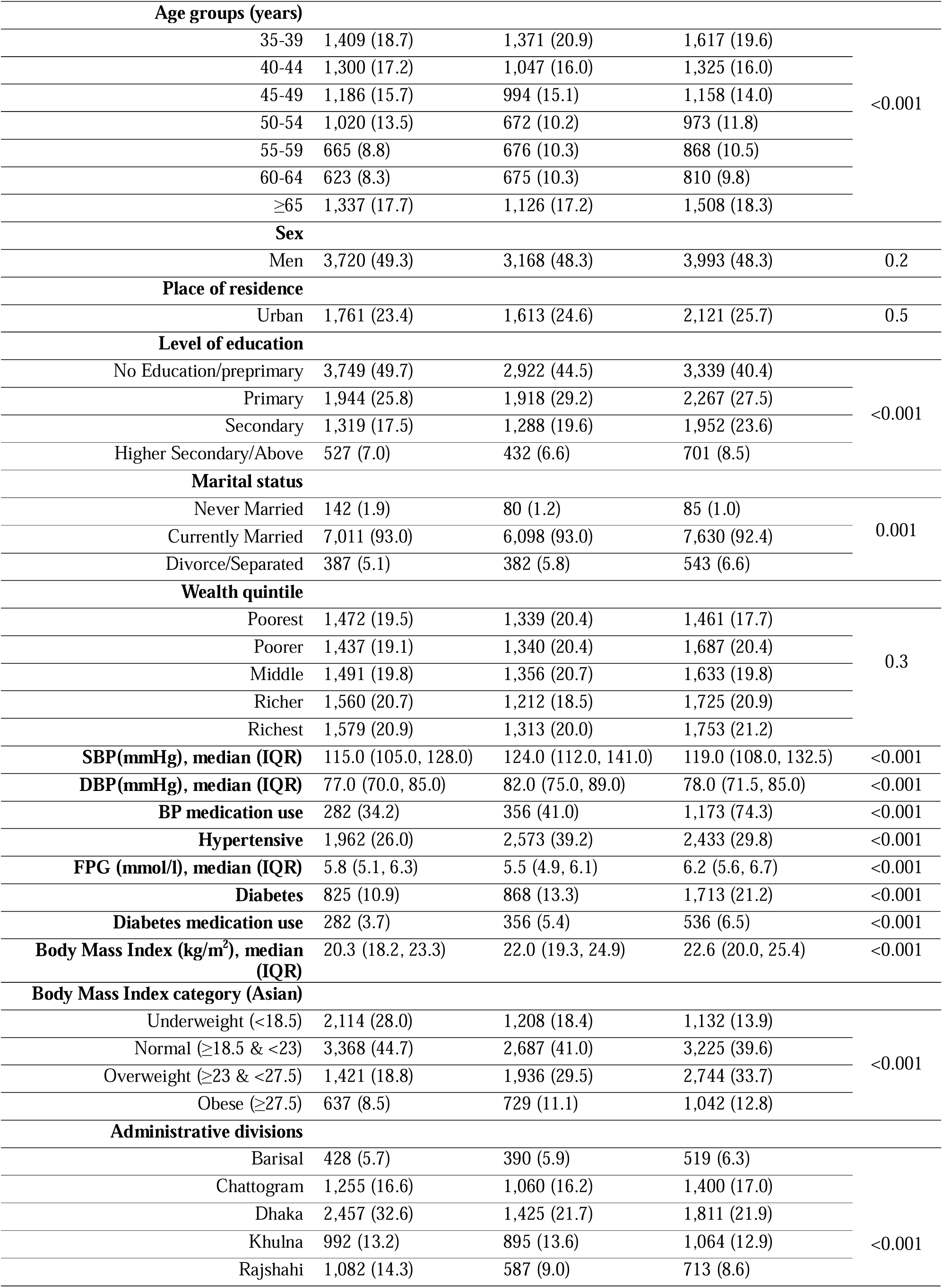
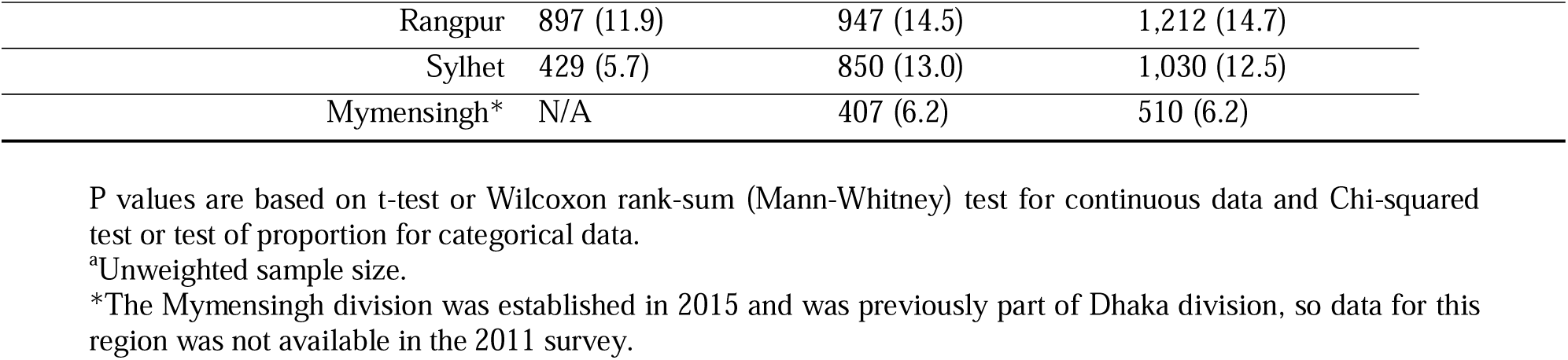
Characteristics of the study population (Weighted) in Bangladesh in 2011-2022.

| Characteristics | Year of Survey |  |  | p-value |
| --- | --- | --- | --- | --- |
|  | 2011 n, % | 2017/2018 n, % | 2022 n, % |  |
| Study sample | 7,540 (7,562<sup>a</sup>) | 6,560 (6, 697<sup>a</sup>) | 8,259 (8,123<sup>a</sup>) |  |
| Age in years, median (IQR) | 49.0 (41.0, 60.0) | 49.0 (40.0, 60.0) | 50.0 (40.0, 60.0) | 0.2 |

| Age groups (years) |  |  |  |  |
| --- | --- | --- | --- | --- |
| 35-39 | 1,409 (18.7) | 1,371 (20.9) | 1,617 (19.6) | <0.001 |
| 40-44 | 1,300 (17.2) | 1,047 (16.0) | 1,325 (16.0) |  |
| 45-49 | 1,186 (15.7) | 994 (15.1) | 1,158 (14.0) |  |
| 50-54 | 1,020 (13.5) | 672 (10.2) | 973 (11.8) |  |
| 55-59 | 665 (8.8) | 676 (10.3) | 868 (10.5) |  |
| 60-64 | 623 (8.3) | 675 (10.3) | 810 (9.8) |  |
| ≥65 | 1,337 (17.7) | 1,126 (17.2) | 1,508 (18.3) |  |
| Sex |  |  |  |  |
| Men | 3,720 (49.3) | 3,168 (48.3) | 3,993 (48.3) | 0.2 |
| Place of residence |  |  |  |  |
| Urban | 1,761 (23.4) | 1,613 (24.6) | 2,121 (25.7) | 0.5 |
| Level of education |  |  |  |  |
| No Education/preprimary | 3,749 (49.7) | 2,922 (44.5) | 3,339 (40.4) | <0.001 |
| Primary | 1,944 (25.8) | 1,918 (29.2) | 2,267 (27.5) |  |
| Secondary | 1,319 (17.5) | 1,288 (19.6) | 1,952 (23.6) |  |
| Higher Secondary/Above | 527 (7.0) | 432 (6.6) | 701 (8.5) |  |
| Marital status |  |  |  |  |
| Never Married | 142 (1.9) | 80 (1.2) | 85 (1.0) | 0.001 |
| Currently Married | 7,011 (93.0) | 6,098 (93.0) | 7,630 (92.4) |  |
| Divorce/Separated | 387 (5.1) | 382 (5.8) | 543 (6.6) |  |
| Wealth quintile |  |  |  |  |
| Poorest | 1,472 (19.5) | 1,339 (20.4) | 1,461 (17.7) | 0.3 |
| Poorer | 1,437 (19.1) | 1,340 (20.4) | 1,687 (20.4) |  |
| Middle | 1,491 (19.8) | 1,356 (20.7) | 1,633 (19.8) |  |
| Richer | 1,560 (20.7) | 1,212 (18.5) | 1,725 (20.9) |  |
| Richest | 1,579 (20.9) | 1,313 (20.0) | 1,753 (21.2) |  |
| SBP(mmHg), median (IQR) | 115.0 (105.0, 128.0) | 124.0 (112.0, 141.0) | 119.0 (108.0, 132.5) | <0.001 |
| DBP(mmHg), median (IQR) | 77.0 (70.0, 85.0) | 82.0 (75.0, 89.0) | 78.0 (71.5, 85.0) | <0.001 |
| BP medication use | 282 (34.2) | 356 (41.0) | 1,173 (74.3) | <0.001 |
| Hypertensive | 1,962 (26.0) | 2,573 (39.2) | 2,433 (29.8) | <0.001 |
| FPG (mmol/l), median (IQR) | 5.8 (5.1, 6.3) | 5.5 (4.9, 6.1) | 6.2 (5.6, 6.7) | <0.001 |
| Diabetes | 825 (10.9) | 868 (13.3) | 1,713 (21.2) | <0.001 |
| Diabetes medication use | 282 (3.7) | 356 (5.4) | 536 (6.5) | <0.001 |
| Body Mass Index (kg/m<sup>2</sup>), median (IQR) | 20.3 (18.2, 23.3) | 22.0 (19.3, 24.9) | 22.6 (20.0, 25.4) | <0.001 |
| Body Mass Index category (Asian) |  |  |  |  |
| Underweight (<18.5) | 2,114 (28.0) | 1,208 (18.4) | 1,132 (13.9) | <0.001 |
| Normal (≥18.5 & <23) | 3,368 (44.7) | 2,687 (41.0) | 3,225 (39.6) |  |
| Overweight (≥23 & <27.5) | 1,421 (18.8) | 1,936 (29.5) | 2,744 (33.7) |  |
| Obese (≥27.5) | 637 (8.5) | 729 (11.1) | 1,042 (12.8) |  |
| Administrative divisions |  |  |  |  |
| Barisal | 428 (5.7) | 390 (5.9) | 519 (6.3) | <0.001 |
| Chattogram | 1,255 (16.6) | 1,060 (16.2) | 1,400 (17.0) |  |
| Dhaka | 2,457 (32.6) | 1,425 (21.7) | 1,811 (21.9) |  |
| Khulna | 992 (13.2) | 895 (13.6) | 1,064 (12.9) |  |
| Rajshahi | 1,082 (14.3) | 587 (9.0) | 713 (8.6) |  |

|  |  |  |  |
| --- | --- | --- | --- |
| Rangpur | 897 (11.9) | 947 (14.5) | 1,212 (14.7) |
| Sylhet | 429 (5.7) | 850 (13.0) | 1,030 (12.5) |
| Mymensingh* | N/A | 407 (6.2) | 510 (6.2) |

P values are based on t-test or Wilcoxon rank-sum (Mann-Whitney) test for continuous data and Chi-squared test or test of proportion for categorical data.
\*Unweighted sample size.
\*The Mymensingh division was established in 2015 and was previously part of Dhaka division, so data for this region was not available in the 2011 survey.

### Age standardized prevalence of diabetes

Table 2 presents the age-standardized prevalence and the relative percentage change of diabetes by risk factors across the BDHS cycles in 2011, 2017/18, and 2022. The overall and sex-specific prevalence of diabetes increased over the survey cycles. The overall prevalence of diabetes was 10.7% (95% CI: 9.8, 11.6) in 2011, 12.8% (95% CI: 11.7, 14.0) in 2017/18, and 20.4% (95% CI: 19.2, 21.7) in 2022, with the highest relative increase of 90.7% in 2022. Between 2011, 2017/18, and 2022, the prevalence of diabetes increased across all age groups; the highest relative increase was seen between 2011 and 2022. The prevalence of diabetes in 2022 was 18.7% (95% CI: 17.3, 20.2) in men and 22.0% (95% CI: 20.4, 23.7) in women, representing a higher relative increase of 96.4% in women than 85.1% in men since 2011. The prevalence of diabetes also increased across all education levels, with the highest relative increase among those with no education (114.5%). The prevalence of diabetes increased substantially between 2011 and 2022 among participants with overweight (45.2%) and obesity (85.7%). All administrative divisions showed a notable increase in diabetes prevalence, with the highest increase in Khulna (168.6%) and Dhaka (154.5%).

**Table 2:** Age standardized prevalence of diabetes in Bangladeshi adults aged 35 years and older, BDHS 2011, 2018 and 2022.

| Characteristics | Age Standardized Prevalence of Diabetes |  |  | Relative % Change |  |
| --- | --- | --- | --- | --- | --- |
|  | 2011 (95% CI) | 2017/18 (95% CI) | 2022 (95% CI) | (2017/18-2011) | (2022-2011) |
| **Overall** | 10.7 (9.8, 11.6) | 12.8 (11.7, 14.0) | 20.4 (19.2, 21.7) | 19.6 | 90.7 |
| **Age groups (years)** |  |  |  |  |  |
| 35-39 | 9.4 (7.7, 11.4) | 11.3 (9.4, 13.4) | 14.6 (12.6, 16.8) | 20.2 | 55.3 |
| 40-44 | 10.7 (8.9, 12.8) | 11.6 (9.6, 14.1) | 18.6 (16.2, 21.2) | 8.4 | 73.8 |
| 45-49 | 10.9 (9.1, 12.9) | 13.1 (10.8, 15.8) | 23.7 (21.0, 26.8) | 20.2 | 117.4 |
| 50-54 | 11.4 (9.4, 13.8) | 16.1 (13.1, 19.5) | 24.2 (21.0, 27.6) | 41.2 | 112.3 |
| 55-59 | 13.6 (11.0, 16.7) | 14.9 (12.2, 18.1) | 25.5 (22.2, 29.1) | 9.6 | 87.5 |
| 60-64 | 12.5 (9.6, 16.1) | 16.7 (13.7, 20.3) | 21.2 (18.1, 24.7) | 33.6 | 69.6 |
| ≥65 | 10.5 (8.8, 12.6) | 12.5 (10.4, 15.0) | 24.1 (21.6, 26.8) | 19.0 | 129.5 |
| **Sex** |  |  |  |  |  |
| Men | 10.1 (9.0, 11.4) | 12.7 (11.3, 14.4) | 18.7 (17.3, 20.2) | 25.7 | 85.1 |
| Women | 11.2 (10.1, 12.5) | 12.8 (11.4, 14.4) | 22.0 (20.4, 23.7) | 14.3 | 96.4 |
| **Place of residence** |  |  |  |  |  |
| Rural | 9.2 (8.2, 10.2) | 11.1 (9.8, 12.6) | 17.2 (16.0, 18.5) | 20.7 | 87.0 |
| Urban | 15.6 (13.6, 17.7) | 17.7 (15.6, 20.1) | 29.6 (26.6, 32.9) | 13.5 | 89.7 |
| **Level of education** |  |  |  |  |  |
| No Education/preprimary | 8.3 (7.2, 9.5) | 10.3 (8.9, 11.9) | 17.8 (16.2, 19.6) | 24.1 | 114.5 |
| Primary | 11.1 (9.5, 12.8) | 12.1 (10.4, 14.0) | 19.5 (17.6, 21.6) | 9.0 | 75.7 |
| Secondary | 12.2 (10.3, 14.3) | 17.0 (14.7, 19.6) | 21.2 (19.2, 23.5) | 39.3 | 73.8 |
| Higher Secondary/Above | 21.5 (18.2, 25.4) | 18.2 (14.6, 22.6) | 31.8 (27.9, 35.8) | -15.3 | 47.9 |
| **Marital status** |  |  |  |  |  |
| Never Married | 8.9 (5.1, 15.3) | 8.4 (4.3, 15.6) | 19.0 (11.2, 30.4) | -5.6 | 113.5 |
| Currently Married | 10.7 (9.8, 11.7) | 12.9 (11.7, 14.2) | 20.3 (19.1, 21.6) | 20.6 | 89.7 |
| Divorce/Separated | 11.3 (8.0, 15.8) | 11.6 (8.3, 15.9) | 22.1 (18.3, 26.4) | 2.7 | 95.6 |
| **Wealth quintile** |  |  |  |  |  |
| Poorest | 7.3 (5.7, 9.3) | 6.7 (5.1, 8.8) | 13.5 (11.5, 15.7) | -8.2 | 84.9 |
| Poorer | 6.9 (5.5, 8.7) | 7.9 (6.3, 9.9) | 16.0 (13.9, 18.3) | 14.5 | 131.9 |
| Middle | 7.6 (6.2, 9.3) | 9.2 (7.3, 11.6) | 18.6 (16.2, 21.2) | 21.1 | 144.7 |
| Richer | 11.1 (9.3, 13.2) | 15.0 (12.5, 17.9) | 23.9 (21.2, 26.8) | 35.1 | 115.3 |
| Richest | 19.7 (17.5, 22.1) | 25.5 (22.7, 28.6) | 29.0 (26.4, 31.7) | 29.4 | 47.2 |
| **Hypertension** | 16.0 (14.1, 18.2) | 17.0 (15.2, 19.0) | 28.0 (25.9, 30.1) | 6.3 | 75.0 |
| **Body Mass Index (kg/m<sup>2</sup>)** |  |  |  |  |  |
| Underweight (<18.5) | 7.9 (6.6, 9.4) | 7.5 (5.9, 9.5) | 11.9 (9.9, 14.2) | -5.1 | 50.6 |
| Normal (≥18.5 & <23) | 8.9 (7.8, 10.3) | 10.1 (8.7, 11.7) | 16.8 (15.1, 18.5) | 13.5 | 88.8 |
| Overweight (≥23 & <27.5) | 15.5 (13.3, 17.9) | 15.8 (13.9, 18.1) | 22.5 (20.6, 24.5) | 1.9 | 45.2 |
| Obese (≥27.5) | 18.2 (14.8, 22.2) | 22.9 (19.3, 27.0) | 33.8 (30.4, 37.4) | 25.8 | 85.7 |
| **Administrative divisions** |  |  |  |  |  |
| Barisal | 12.2 (10.0, 14.7) | 11.6 (8.6, 15.5) | 17.7 (14.9, 20.8) | -4.9 | 45.1 |
| Chattogram | 13.9 (11.7, 16.6) | 15.4 (12.3, 19.2) | 23.4 (20.7, 26.4) | 10.8 | 68.3 |
| Dhaka | 11.0 (9.1, 13.3) | 19.5 (16.2, 23.3) | 28.0 (24.3, 32.1) | 77.3 | 154.5 |
| Khulna | 7.0 (5.7, 8.6) | 10.3 (8.4, 12.7) | 18.8 (16.2, 21.7) | 47.1 | 168.6 |
| Rajshahi | 10.4 (8.6, 12.6) | 10.3 (8.0, 13.1) | 14.0 (11.3, 17.2) | -1.0 | 34.6 |
| Rangpur | 8.5 (6.5, 11.0) | 9.5 (7.2, 12.5) | 13.6 (11.3, 16.3) | 11.8 | 60.0 |
| Sylhet | 11.5 (9.6, 13.9) | 7.5 (5.6, 10.0) | 18.2 (15.1, 21.7) | -34.8 | 58.3 |
| Mymensingh | N/A | 10.7 (7.7, 14.8) | 20.9 (17.3, 25.1) | N/A | N/A |

Data are presented as percentages with 95% confidence intervals. Prevalence estimates are age-standardized to the population of individuals aged 35 years and older, based on the 2022 Bangladesh Population and Housing Census.

### Increasing trends in diabetes prevalence and fasting plasma glucose

Figure 1 presents the PR of diabetes and the mean difference of FPG among Bangladeshi adults across the BDHS cycles, adjusted for covariates. Overall, the prevalence of diabetes increased by 37% (PR: 1.37; 95% CI: 1.31, 1.43) with each subsequent BDHS cycle compared to the BDHS 2011 cycle. Men had a PR of 1.28 (95% CI: 1.20, 1.37), while women exhibited the highest increase, with a PR of 1.43 (95% CI: 1.34, 1.52) per cycle.

**Figure 1:**
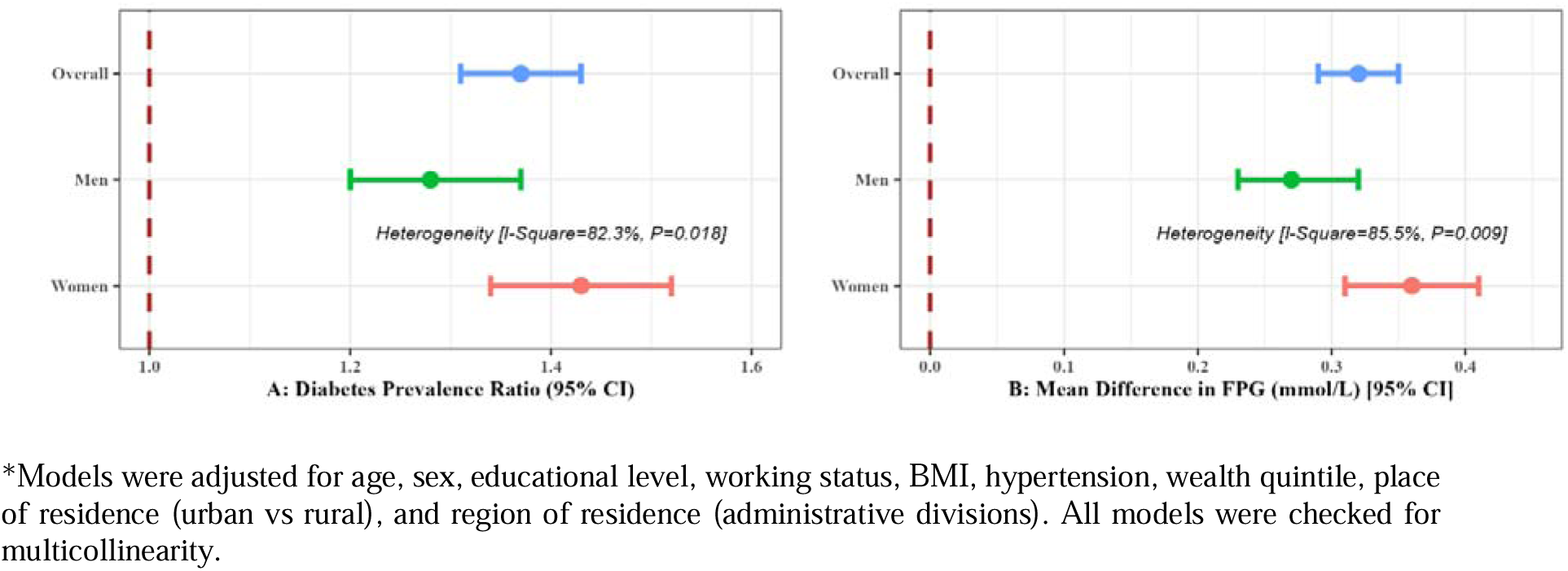
The prevalence ratio of diabetes and the mean difference of fasting plasma glucose (mmol/L) among Bangladeshi adults by the BDHS cycles

For FPG levels, the mean difference indicated an overall increase of 0.32 mmol/L (95% CI: 0.29, 0.35), with men experiencing an increase of 0.27 mmol/L (95% CI: 0.23, 0.32) and women showing a larger increase of 0.36 mmol/L (95% CI: 0.31, 0.41).

The increasing trends in FPG are consistent with PRs. A heterogeneity test confirmed that the upward trends were significantly more pronounced in women than in men. Notably, the changes in diabetes prevalence and FPG were substantially higher between 2017/18 and 2022, suggesting a non-linear pattern across the BDHS cycles (Supplementary Figure 1).

### Risk factors for diabetes and fasting plasma glucose

Table 3 highlights the risk factors associated with the changes in diabetes and FPG adjusted for BDHS cycles. The prevalence of diabetes increased across all age groups compared to the younger population, with the highest PR observed in the 55 to 59 years age group (PR: 1.62, 95% CI: 1.42, 1.85). Similarly, diabetes prevalence rose across all BMI categories, education levels, and wealth quintiles. The highest PRs were observed among individuals with obesity (PR: 1.80, 95% CI: 1.57, 2.07), those with the highest education level (PR: 1.32, 95% CI: 1.17, 1.49), and those in the highest wealth quintile (PR: 1.78, 95% CI: 1.57, 2.02), when compared to individuals with underweight, no formal education, and in the lowest wealth quintile, respectively. Additionally, people residing in urban areas (PR: 1.26, 95% CI: 1.17, 1.36) and those living with hypertension (PR: 1.33, 95% CI: 1.24, 1.42) exhibited higher PRs compared to those living in rural areas and those without hypertension. The risk factors for diabetes were identical in both men and women. The trend and the risk factors for FPG were consistent across both the overall population and sex-specific analyses (see Table 3).

**Table 3:**
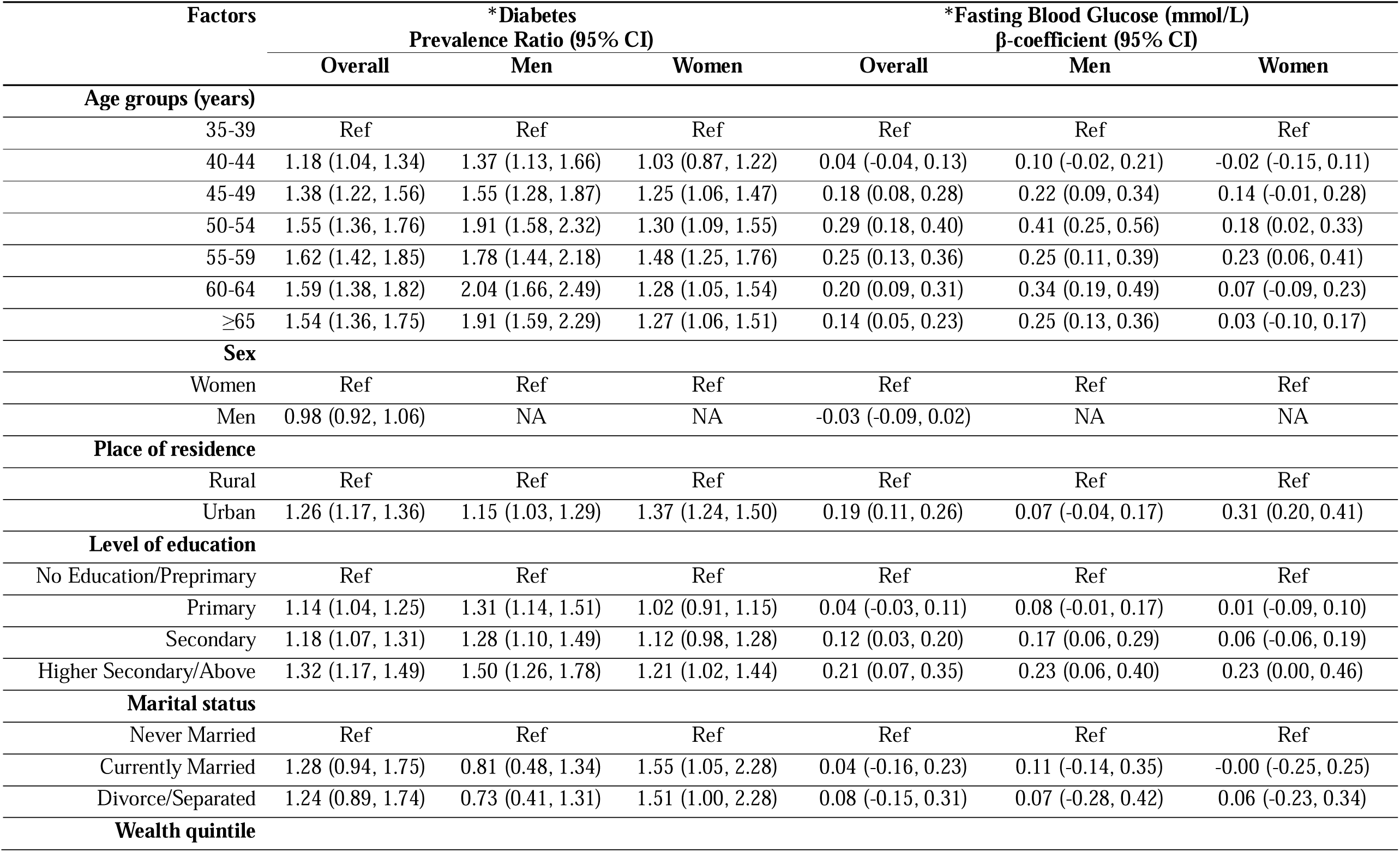
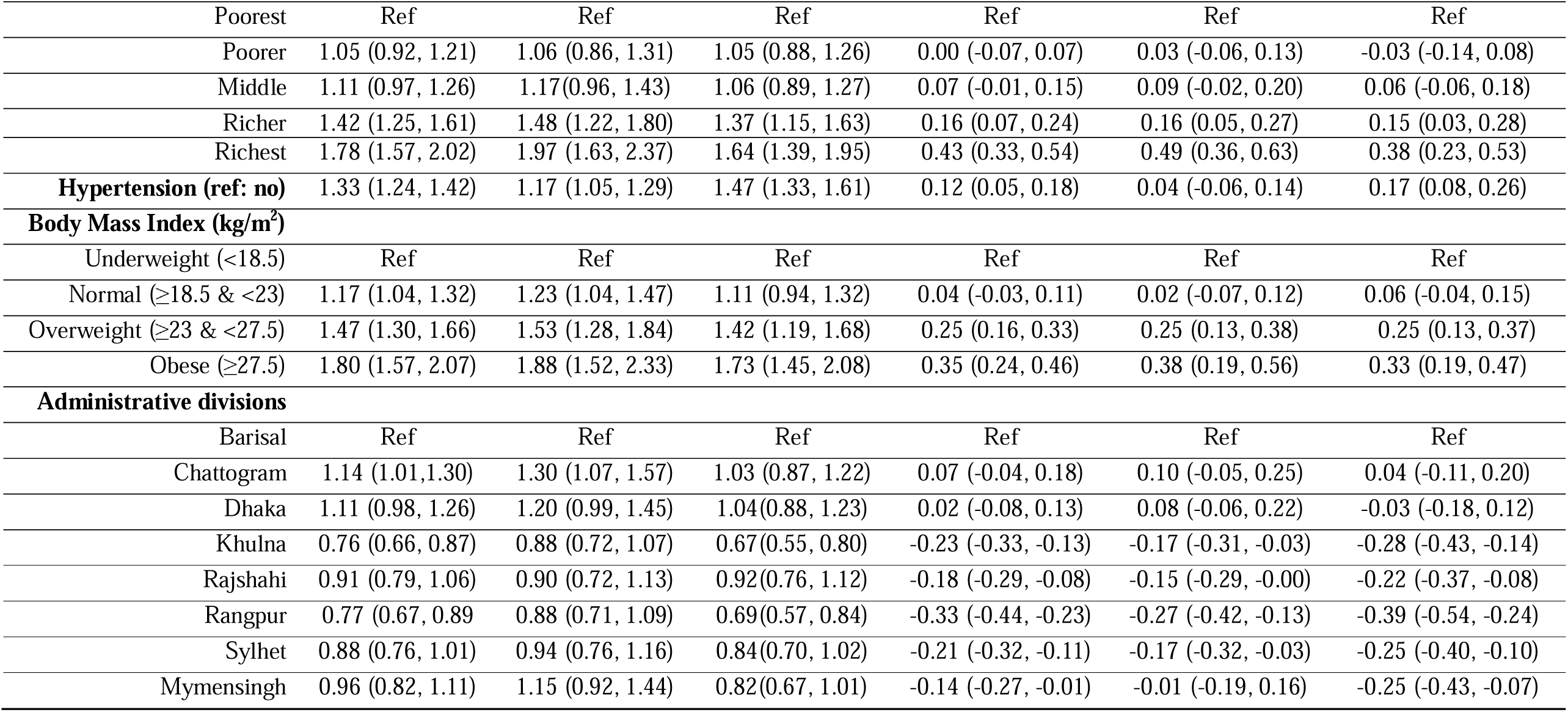
Risk factors for changes in diabetes and fasting plasma glucose among Bangladeshi adults aged 35 years and older.

## Discussion

Our analyses demonstrated a consistent increase in both overall and sex-specific diabetes prevalence and FPG between 2011 and 2022. The overall age-standardised prevalence of diabetes was 10.7% (95% CI: 9.8, 11.6) in 2011, 12.8% (95% CI: 11.7, 14.0) in 2017/18, and 20.4% (95% CI: 19.2, 21.7) in 2022, with the highest relative increase of 90.7% observed by 2022. The PRs increased by 37% per BDHS cycle, with women showing the largest increase at 43%. Likewise, the overall FPG increased by 0.32 mmol/L with a greater rise in women (0.36 mmol/L) compared to men (0.27 mmol/L). These increases followed a non-linear pattern over time, with BMI and socioeconomic status emerging as major risk factors for changes in diabetes prevalence and FPG levels. The findings of this study underscore the urgent need for population-level interventions to control this increasing burden of diabetes in Bangladesh, as the age-standardized prevalence of diabetes and FPG levels have both risen across all risk factors.

This steeper rise in diabetes prevalence among women may be due to multiple factors. Cultural and social norms in Bangladesh often limit physical activity options for women, contributing to higher levels of inactivity—a known diabetes risk factor. A recent scoping review in Bangladesh revealed that the prevalence of insufficient physical activity was notably higher in women, ranging from 27% to 54%, compared to men, whose prevalence ranged from 7% to 34% ^22^. Women face unique risks that increase insulin resistance, including inadequate preconceptual care, gestational diabetes and hormonal changes around menopause ^23^, which may contribute to heightened diabetes susceptibility among women in Bangladesh and warrant further investigation. Moreover, during the early phases of Bangladesh’s nutrition-epidemiological transition, women may be disproportionately affected by shifts toward high-calorie, processed foods. For example, overweight and obesity rates have increased significantly in women from 2011 to 2022 (p < 0.001, data not shown). Socioeconomic barriers in Bangladesh, including financial and educational limitations, further restrict women’s access to preventive care and lifestyle guidance, exacerbating health risks.

Our findings are consistent with other studies that have found secular increases in the prevalence of diabetes in Bangladesh ^24^. A scoping review of 22 studies between 1994 and 2013 found an increasing trend of diabetes prevalence in Bangladesh at a rate of between 0.05% and 0.06% per year ^24^. However, many of the included studies were not nationally representative, and/or sampled participants with prediabetes (FPG in the range 6.1 to 6.9 mmol/L) ^13^, artificially inflating diabetes prevalence estimates. Our analysis provides more robust evidence of an increasing trend in diabetes and FPG in Bangladesh over the last decade, with a notably higher increase among women compared to men.

Our findings are also consistent with global projections indicating a rise in diabetes prevalence worldwide by 2045, with most of the burden expected in LMICs ^2,25,26^. While some high-income countries (HICs) have implemented diabetes prevention strategies in primary care settings ^27,28^, broader adoption is needed in LMICs. Among high-risk individuals, diabetes prevalence rose by approximately 50–80% across BDHS cycles, a worrying trend that, if sustained, will impose a substantial diabetes burden on Bangladesh. This pattern reflects secular increases in obesity observed both in Bangladesh ^29^ and globally^30^, likely due to shifts toward energy-dense diets and reduced physical activity ^31^.

We found that diabetes prevalence increased most among individuals in the richer and richest socioeconomic groups, as well as among those with higher education levels, compared to the poorest and least educated individuals. These findings are consistent with Bangladesh being in the early stages of a nutrition-epidemiological transition. This transition often begins in LMICs with an increase in sedentary behaviour and the consumption of high-energy foods, leading to rising rates of obesity, hypertension, and diabetes ^32^. In later stages, higher socioeconomic groups tend to adopt healthier lifestyles, including physical activity and balanced diets, as observed in HICs ^33^. In our study, the increase in BMI and diabetes prevalence among the ‘rich’ and ‘educated’ groups suggests that Bangladesh is in the early phases of this transition. As economic development continues, the burden of diabetes will continue to shift to lower socioeconomic groups, as seen in HICs ^34^.

The rising prevalence of diabetes in Bangladesh is likely due to the lack of effective preventive interventions. Although the Government of Bangladesh has introduced the Multisectoral Action Plan for the Prevention and Control of Noncommunicable Diseases (NCDs) (2018–2025), aligned with WHO’s Package of Essential NCD Interventions for primary care, the approach remains primarily curative and management-focused, applied mainly at Upazila Health Complexes, with screening limited to Community Clinics ^35^. Despite the plan’s goal to halt the rise in obesity and diabetes by 2025, community-level preventive strategies essential for reducing the diabetes burden are largely absent.

The implications of these findings are that there needs to be a stronger focus on diabetes prevention directed at both individual and population levels as our data suggests substantial disparities in these factors. Our findings suggest that tackling the rising rates of overweight and obesity will be crucial for any effective diabetes prevention strategy in Bangladesh. For example, population level measures by the government are needed to provide more incentives for purchasing whole grain, nuts and seeds, fruits and vegetables and increasing physical activity, while putting restrictions or disincentives for less healthy products (1). Efforts to prevent diabetes should be population-wide; however, given the stronger association of diabetes with higher wealth quintiles in our findings, targeted interventions for lower wealth quintiles are also crucial to prevent a future shift in the burden. This approach aligns with patterns observed in later stages of the nutrition-epidemiological transition, where lower socioeconomic groups become increasingly affected by diabetes as countries continue to develop. Given the rising number of diabetes cases in LMICs, attention should also be given to health service delivery through early case detection and linkage to care to improve the management of existing cases in Bangladesh. These strategies will help ensure Bangladesh meets the World Health Organization Global Action Plan for the Prevention and Control of NCDs in 2025, to attain a 25% relative reduction in premature mortality from NCDs by 2025 as well as sustainable development goals target of one-third reduction of premature mortality related to NCDs from the 2015’s level by 2030 ^1,36^.

A major strength of this study is that it examined trends in diabetes prevalence using three large, independent, nationally representative surveys, all employing the same methodology. Additionally, diabetes was assessed based on WHO classification criteria. Our use of the GEE model provided robust population-averaged estimates, effectively accounting for within-subject and within-cluster correlations, and produced reliable standard errors despite the complex survey design. However, the study is limited to individuals aged 35 years and older, as the BDHS 2011 did not include younger populations. Furthermore, we were unable to adjust for dietary habits, physical activity, smoking and environmental factors which are key predictors of diabetes, as these data were not collected.

In conclusion, there was a significant increase in the prevalence of diabetes and FPG among Bangladeshi adults from 2011 to 2022, with a particularly sharp rise observed between 2017/18 and 2022, especially among women. BMI and socio-economic status were the key risk factors for changes in diabetes and elevated FPG. Population-level interventions are urgently needed to control this increasing burden of diabetes in Bangladesh.

## Contributors

JR: Conceptualisation, Data curation and analysis, Interpretation of the data, Critical revision of the manuscript and Final approval.

JCO: Interpretation of the data, Writing of the initial draft, Critical revision of the manuscript and Final approval.

MNK, SR: Data curation and analysis, Critical revision of the manuscript and Final approval.

MBH, SK, DM, MNK: Interpretation of the data, Critical revision of the manuscript and Final approval.

RMI: Conceptualisation, Data curation and analysis, Interpretation of the data, Writing of the initial draft, Critical revision of the manuscript and Final approval.

## Data sharing statement

The data underlying this article are available in the manuscript, in online *Supplementary materials* and in our records. These data can be shared on request.

## Declaration of interests

All authors declare that they have no potential conflicts of interest.

## Data Availability

All data produced in the present study are available upon reasonable request to the authors

https://microdata.worldbank.org/index.php/catalog/dhs/?page=1&sk=BDHS&ps=15&repo=dhs

## Acknowledgements

The authors thank to MEASURE DHS for granting access to the BDHS data.

---

**Supplementary Figure 1:**
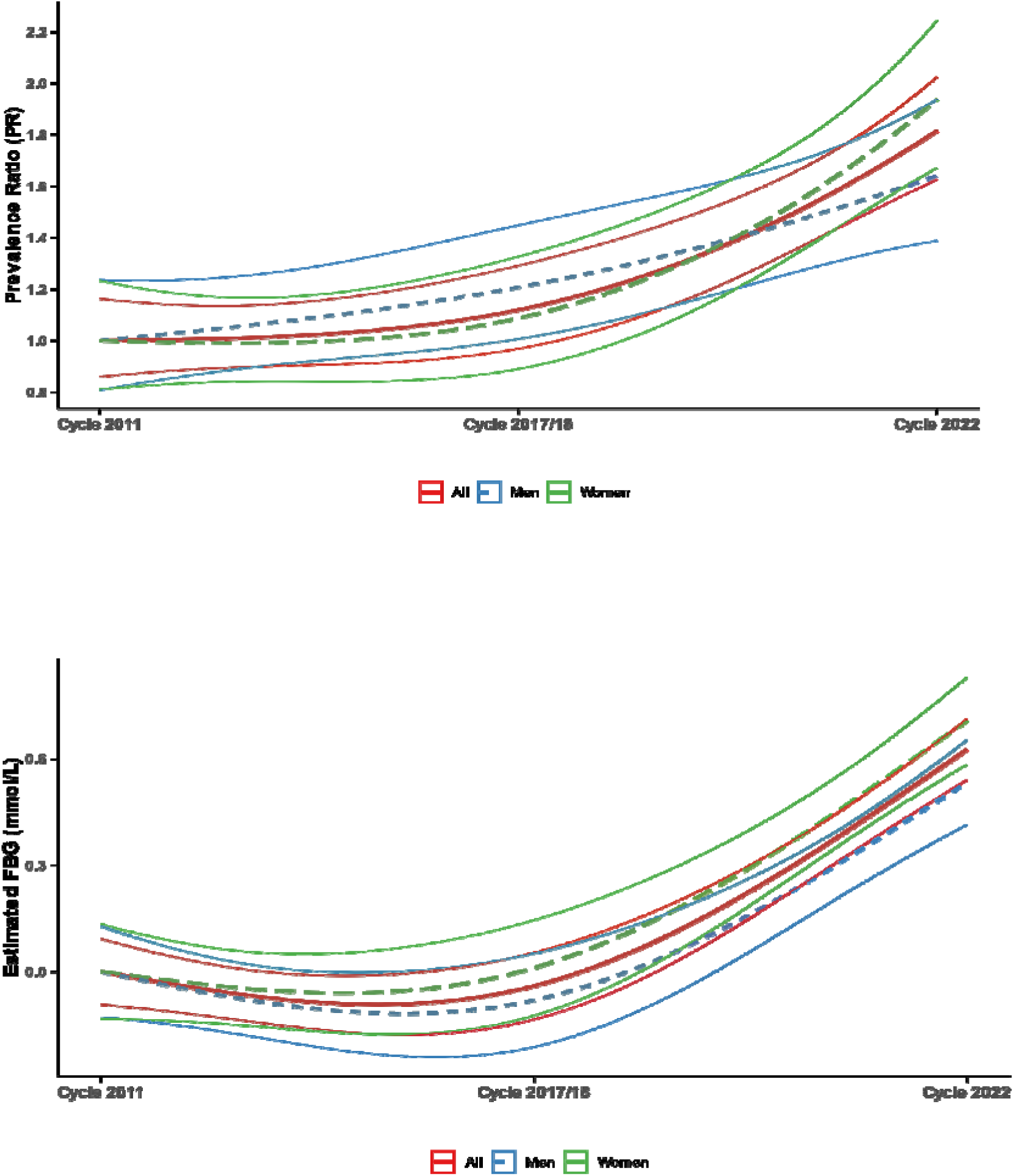
Trends in diabetes prevalence ratio and an estimated fasting plasma glucose (mmol/L) among Bangladeshi adults by the BDHS cycles

## Notes

### Competing Interest Statement

The authors have declared no competing interest.

### Funding Statement

This study did not receive any funding

### Author Declarations

https://microdata.worldbank.org/index.php/catalog/dhs/?page=1&sk=BDHS&ps=15&repo=dhs

## References

1. Lin X, Xu Y, X P, Jingya Xu J, Ding Y, Sun X, … Shan P-F. Global, regional, and national burden and trend of diabetes in 195 countries and territories: an analysis from 1990 to 2025. Nature 2020; 10: 14790.

2. Sun H, Saeedi P, Karuranga S, Pinkepank M, Ogurtsova K, Duncan BB, … Mbanya JC. IDF diabetes atlas: Global, regional and country-level diabetes prevalence estimates for 2021 and projections for 2045. Diabetes research and clinical practice 2021: 109119.

3. International Diabetes Federation. IDF Diabetes Atlas, 10th edn. Brussels. Belgium: 2021. Available at: https://diabetesatlas.org/, accessed 20 January 2022, 2021.

4. Marchionni N, Di Bari M, Fumagalli S, Ferrucci L, Baldereschi G, Timpanelli M, Masotti G. Variable effect of comorbidity on the association of chronic cardiac failure with disability in community-dwelling older persons. Archives of gerontology and geriatrics 1996; 23(3): 283–92.

5. Vollset SE, Goren E, Yuan C-W, Cao J, Smith AE, Hsiao T, … Chalek J. Fertility, mortality, migration, and population scenarios for 195 countries and territories from 2017 to 2100: a forecasting analysis for the Global Burden of Disease Study. The Lancet 2020; 396(10258): 1285–306.

6. Afroz A, Habib SH, Chowdhury HA, Paul D, Shahjahan Md, Hafez A, … Ali L. Healthcare cost of type 2 diabetes mellitus in Bangladesh: a hospital-based study. Int J Diabetes Dev Ctries 2016; 36(2): 235–41.

7. Zimmet PZ, Magliano DJ, Herman WH, Shaw JE. Diabetes: a 21st century challenge. Lancet Diabetes Endocrinol 2014; 2: 56–64.

8. Akter S, Rahman MM, Abeb SK, Sultanac P. Prevalence of diabetes and prediabetes and their risk factors among Bangladeshi adults: a nationwide survey. Bull World Health Organ 2014; 92: 204–13A.

9. National Institute of Population Research and Training (NIPORT) Ma, and ICF International,. Bangladesh Demographic andHealth Survey 2011. Dhaka, Bangladesh and Calverton, Maryland, USA:NIPORT, Mitra and Associates, and ICF International, 2013.

10. Hossain MB, Khan MN, Oldroyd JC, Rana J, Magliago DJ, Chowdhury EK, … Islam RM. Prevalence of, and risk factors for, diabetes and prediabetes in Bangladesh: Evidence from the national survey using a multilevel Poisson regression model with a robust variance. PLOS Global Public Health 2022; 2(6): e0000461.

11. National Institute of Population Research and Training (NIPORT) Ma, and ICF International,. Bangladesh Demographic and Health Survey 2022. Dhaka, Bangladesh and Calverton, Maryland, USA:NIPORT, Mitra and Associates, and ICF International, 2024.

12. National Institute of Population Research and Training (NIPORT) Ma, and ICF International,. Bangladesh Demographic andHealth Survey 2017-2018. Dhaka, Bangladesh and Calverton, Maryland, USA:NIPORT, Mitra and Associates, and ICF International, 2020.

13. World Health Organization. Definition and diagnosis of diabetes mellitus and intermediate hyperglycaemia: report of a WHO/IDF consultation. 2006.

14. Islam RM, Magliano DJ, Khan MN, Hossain MB, Rana J, Oldroyd JC. Prevalence of undiagnosed diabetes and the relative importance of its risk factors among adults in Bangladesh: findings from a nationwide survey. Diabetes Research and Clinical Practice 2022; 185: 109228.

15. Anjana RM, Deepa M, Pradeepa R, Mahanta J, Narain K, Das HK, … Kumar A. Prevalence of diabetes and prediabetes in 15 states of India: results from the ICMR–INDIAB population-based cross-sectional study. The lancet Diabetes & endocrinology 2017; 5(8): 585–96.

16. Basit A, Fawwad A, Qureshi H, Shera A. Prevalence of diabetes, pre-diabetes and associated risk factors: second National Diabetes Survey of Pakistan (NDSP), 2016–2017. BMJ open 2018; 8(8): e020961.

17. Rutsein SO, Johnson K. The DHS Wealth Index. DHS Comparative Reports No. 6.. Calverton, Maryland, USA: ORC Macro. Available at http://dhsprogram.com/publications/publication-cr6-comparative-reports.cfm. Accessed November 07, 2015., 2004.

18. Barros AJ, Hirakata VN. Alternatives for logistic regression in cross-sectional studies: an empirical comparison of models that directly estimate the prevalence ratio. BMC medical research methodology 2003; 3(1): 21.

19. Hanley JA, Negassa A, Edwardes MDd, Forrester JE. Statistical analysis of correlated data using generalized estimating equations: an orientation. American journal of epidemiology 2003; 157(4): 364–75.

20. Kaufman JS, MacLehose RF. Which of these things is not like the others? Cancer 2013; 119(24): 4216–22.

21. Von Elm E, Altman DG, Egger M, Pocock SJ, Gøtzsche PC, Vandenbroucke JP. The Strengthening the Reporting of Observational Studies in Epidemiology (STROBE) statement: guidelines for reporting observational studies. Annals of internal medicine 2007; 147(8): 573–7.

22. Uddin R, Hasan M, Saif-Ur-Rahman K, Mandic S, Khan A. Physical activity and sedentary behaviour in Bangladesh: a systematic scoping review. Public Health 2020; 179: 147–59.

23. Kautzky-Willer A, Leutner M, Harreiter J. Sex differences in type 2 diabetes. Diabetologia 2023; 66(6): 986–1002.

24. Biswas T, Islam A, Rawal LB, Islam S. Increasing prevalence of diabetes in Bangladesh: a scoping review. Public health 2016; 138: 4–11.

25. Guariguata L, Whiting DR, Hambleton I, et al. Global estimates of diabetes prevalence for 2013 and projections for 2035.. Diabetes Res Clin Pract 2014; 103: 137–49.

26. Gujral UP, Pradeepa R, Weber MB, Venkat Narayan KM, Mohan V. Type 2 diabetes in South Asians: similarities and differences with white Caucasian and other populations. Ann NY Acad Sci 2013; 1281: 51–63.

27. Oldenburg B, Absetz P, Dunbar JA, Reddy P, O’Neil A. The spread and uptake of diabetes prevention programs around the world: a case study from Finland and Australia. Transl Behav Med 2011; 1(2): 270–82.

28. Centre for Disease Control and Prevention. National Diabetes Prevention Program https://www.cdc.gov/diabetes/prevention/index.html Accessed 21st November 2021. 2021.

29. Chowdhury MAB, Adnan MM, Hassan MZ. Trends, prevalence and risk factors of overweight and obesity among women of reproductive age in Bangladesh: a pooled analysis of five national cross-sectional surveys. BMJ open 2018; 8(7): e018468.

30. Ng M, Fleming T, Robinson M, et al. Global, regional, and national prevalence of overweight and obesity in children and adults during 1980-2013: a systematic analysis for the Global Burden of Disease Study 2013. The Lancet 2014; 384: 766–81.

31. Malik VS, Willett WC, Hu FB. Global obesity: trends, risk factors and policy implications. Nature reviews endocrinology 2013; 9(1): 13–27.

32. Jaacks LM, Vandevijvere S, Pan A, McGowan CJ, Wallace C, Imamura F, … Ezzati M. The obesity transition: stages of the global epidemic.. Lancet Diabetes Endocrinol 2019; 7: 231–40.

33. Popkin BM. An overview on the nutrition transition and its health implications: the Bellagio meeting.. Public health nutrition 2002; 5: 93–103.

34. Deepa M, Anjana RM, Manjula D, Narayan KMV, Mohan V. Convergence of prevalence rates of diabetes and cardiometabolic risk factors in middle and low income groups in urban India: 10-year follow-up of the Chennai Urban Population Study. J Diabetes Sci Technol 2011; 5: 918–27.

35. Riaz BK, Ali L, Ahmad SA, Islam MZ, Ahmed KR, Hossain S. Community clinics in Bangladesh: a unique example of public-private partnership. Heliyon 2020; 6(5).

36. UN General Assembly. Transforming our world: the 2030 agenda for sustainable development. https://sustainabledevelopment.un.org/post2015/transformingourworld/publication, accessed on 17 January 2022. 2015.

